# Assessment of venous thromboembolism risk and use of anticoagulant thromboprophylaxis in CHUK, Rwanda: a cross-sectional study

**DOI:** 10.1101/2022.12.26.22283948

**Authors:** Emile Abimana, Etienne Ntabanganyimana, Raphael Ndahimana, R Osee Sebatunzi, Florence Masaisa

## Abstract

**Background:** Venous thromboembolism (VTE) is common and preventable disease among non-surgical hospitalized patients. Its incidence is high and alarming. Acute medical patients have VTE risk during and after hospitalization. Padua prediction score is a risk model created to identify high VTE risk patients among non-surgical hospitalized patients.

**Methods:** We performed a cross-sectional survey of 107 patients admitted in Internal Medicine wards that were assessed as acute ill patients at Emergency Department, in a period of 4 weeks. The demographic and clinical data were collected using a designed questionnaire. VTE risk was defined as having a Padua Prediction score of ≥4 points. Statistical analysis was done to determine prevalence. The patients at high VTE risk received thromboprophylaxis.

**Results:** Of one hundred and seven eligible patients included. 84% were found with high VTE risk. Among physiologically unstable patients, 60% of the patients were classified in red color during the time of triage, this means, they were severely sick and needing resuscitation. Among leading diagnosis, severe pneumonia was predominant (29%). Severe pneumonia and uncontrolled DM showed significant association with high VTE risk. 11.1% of high VTE risk patients were taking anticoagulant thromboprophylaxis prior to the recruitment.

**Conclusion:** This study demonstrated a high prevalence of VTE risk among acute ill medical patients and underuse of anticoagulants thromboprophylaxis in potential patients at Kigali University Teaching Hospital, CHUK. Padua prediction score should be implemented for early detection of patients at-risk of VTE in severely ill patients and start anticoagulant thromboprophylaxis on time for reducing mortality and morbidity.

## BACKGROUND

Venous thromboembolism (VTE), that includes deep venous thrombosis (DVT) and pulmonary embolism (PE), is one of the most causes of morbidity and mortality in acutely ill and critical medical or surgical inpatients [1]. In non-surgical patients, the incidence is ranging between 10 to 30% and associated with 10% of mortality rate, three fourth of deaths occur in medical patients [2,3]. The use of VTE prophylaxis decreases its risk in surgical patients besides high risk of bleeding due to the multiple comorbidities [1]. The VTE was identified to be among the preventable diseases in order to decrease mortality and morbidity, in 2008, in US, they have introduced the VTE prophylaxis protocols [4,5]. Risks for VTE include immobility, infections, heart failure, respiratory failure, and active or underlying malignancy; it includes also increase of age, prior VTE and underlying congenital or acquired thrombophilia, these risks are synergistic and the more risk factors present, the higher the VTE risk [6]. The guidelines by the American College of Chest Physician (ACCP) recommends the use of anticoagulant thromboprophylaxis with low molecular weight heparin, or low dose unfractionated heparin or fondaparinux for medical patients at risk of thrombosis, the VTE risk evaluation done by using Padua prediction score and discourages the use of anticoagulants in medical patients with active bleeding or at high risk of bleeding [7]. Despite the ACCP recommendation, globally, a half of hospitalized medical patients with a high risk of DVT received thromboprophylaxis [7,8]. There is a risk assessment model which was predefined by the Padua Prediction Score to assign eleven common VTE risk in medical patients, the score of ≥4 points defined as high risk and this model provided the best assessment basis but with some weakness, there was establishment of a bleeding risk score besides not validated and no separation of use versus nonuse of prophylaxis [7,9]. There major bleeding risks such as: bleeding within 3 months prior to admission, platelets count less than 50×10^9^/L and active gastro-duodenal bleeding [7,10]. In CHUK, Emergency Department, they are using modified South Africa Triage Score (mSATS) during the triage where the patients are classified into five colors (red, orange, yellow, green and blue) according to the clinical status [10]. Unstable patients requiring resuscitation are labeled red, yellow and green patients are not severely sick while blue patients are clearly dead [11]. Many interventions may improve the overall compliance of VTE risk assessment in acute medical ill patients and results in a positive attitude towards mortality and morbidity reduction, improvement of adherence to pharmacology thromboprophylaxis as preventive measures, the advocacy of VTE risk assessment is paramount at all health facilities level [11]. In absence of standard protocol, this study was done to see if all severely ill medical patients at-risk of VTE at CHUK are identified and put on anticoagulant thromboprophylaxis, we assessed the magnitude of VTE risk and the use of anticoagulant thromboprophylaxis in acutely ill medical patients in CHUK, Rwanda.

## METHODS

### Study design and methodology

Kigali University Teaching Hospital, CHUK is a public, tertiary hospital in Rwanda, located in Kigali, Capital City of Rwanda. It is the biggest hospital among three major referral hospitals located in Kigali with 560 beds capacity. The Internal Medicine is among the departments based on type of specialties. We performed a non-randomized, cross-sectional, descriptive and observational study. During 4 weeks, between April and May, 2020; 107 patients aged ≥ 18 years old, who were admitted in Internal Medicine through emergency rooms, triaged in red (patients who were physiologically unstable and required resuscitation) or orange (patients who were potentially unstable physiologically or had life threatening pathology) and accepted or the legal next of kin accepted to give consent, were recruited in the study. At Emergency Department, the triage is done by using a local modified South Africa Triage Score (mSATS).The patients who had active bleeding, gastro duodenal ulcer, platelets count <50×10^9^/L, known with bleeding risks (like hemophilia or bleeding within 3 months prior to admission), and or incomplete data in their chart from Emergency department were excluded from the study. The data collection was done using a tool of variables; the Padua Prediction Score (Table 1), overall the cumulative score of ≥4 points was recorded as high VTE risk and the patients had benefited anticoagulant thromboprophylaxis. The obesity (BMI≥30) was not assessed; many patients were severely sick and bedridden.

**Table 1.** Padua prediction score assessment tool: High risk VTE score: ≥ 4 points. Source: Dr. Sofia Barbar at MD+CALC.

| Item(s) | score |
| --- | --- |
| **Active cancer** | 3 |
| **Previous VTE** | 3 |
| **Bed rest for ≥3 days** | 3 |
| **Thrombophilia** | 3 |
| **Recent trauma (≤ 1 month)** | 2 |
| **Elderly age (≥70 years)** | 1 |
| **Heart failure/Respiratory failure** | 1 |
| **Acute myocardial infection/ischemic stroke** | 1 |
| **BMI ≥30 Kg/m<sup>2</sup>** | 1 |
| **Ongoing hormonal therapy** | 1 |

### ANALYSIS

Date entry was done using Epidata version 3.1 and exported to excel and the statistical package for social science (IBM SPSS Statistics) version 28 for final data cleaning and analysis. We performed descriptive statistics determine frequency and percentages and later we did chi-square test of independence and Binary logistic regression to find out the relevant significance of association where by p-value <0.05 under confidence of 95% the predictive variable(s) was considered as significantly associated with outcome variable. No confounding variables identified in literature review fitting for multinomial logistic regression testing for adjustment.

## RESULTS

Of 107 patients, almost 59.8% were female, 60.7% were classified in red color in triage at the time of admission at accident and emergency department, among leading diagnosis one three had severe pneumonia, 84.1% had high VTE risk (Table 2). In assessment of venous thromboembolism by Padua prediction score, 86.9% did not report active bleeding, 95.3% denied history of thrombophilic condition, 91.6% were active cancer free, 91.6% were not in heart failure, 80% were younger than 70 years old, 89.7% did not have ischemic stroke and 71% did not have acute infection (Table 3). During bivariate analysis of the leading diagnosis and Padua prediction score, severe pneumonia diagnosis had significant association with venous thromboembolism (p value = 0.004) and the same as uncontrolled Diabetes Mellitus with p value < 0.001 (Table 4). Among the 10 patients who were taking anticoagulant thromboprophylaxis, 3 patients had severe pneumonia (Table 5) and there was no significant association between Padua prediction score and VTE prophylaxis (p value = 0.234) (Table 6). In Padua prediction score distribution, in study population, 62.6% scored 4 points, 7.5% scored the highest score in this study which was 6 points, and 15.9% scored less than 4 points means no VTE risk (Figure 1). In Padua prediction score distribution by mean age, the patients with 77.3 years old scored 5 points and patients with 53.4 years old scored 6 points (Figure 2). Blindly, 11.1% of high VTE risk patients were taking anticoagulant thromboprophylaxis prior to our study recruitment (Table 3)

**Table 2.** Baseline characteristics of the patients enrolled Source: Authors’ Compilation, 2020

| Characteristics | Number(n) | Percentage(%) |
| --- | --- | --- |
| **Gender** |  |  |
| **Male** | 43 | 40.2 |
| **Female** | 64 | 59.8 |
| **Total** | 107 | 100 |
| **Triage color of patient** |  |  |
| **RED** | 65 | 60.7 |
| **ORANGE** | 42 | 39.3 |
| **Total** | 107 | 100 |
| Leading Diagnosis |  |  |
| **Acute decompensated heart failure** | 9 | 8.4 |
| **complicated malaria** | 11 | 10.3 |
| **Severe pneumonia** | 31 | 29.0 |
| **Extra-pulmonary TB** | 6 | 5.6 |
| **Ischemic stroke** | 11 | 10.3 |
| **Malignancy** | 9 | 8.4 |
| **DKA** | 9 | 8.4 |
| **Meningitis** | 8 | 7.5 |
| **uncontrolled DM** | 13 | 12.1 |
| **Total** | 107 | 100.0 |
| Padua score |  |  |
| **<4** | 17 | 15.9 |
| **≥4** | 90 | 84.1 |
| **Total** | 107 | 100 |

**Table 3.** Descriptive statistics of Padua prediction score Elements Source: Authors’ Compilation, 2020

| Characteristics | Number(s) | Percentage(s) |
| --- | --- | --- |
| Active bleeding |  |  |
| **Yes** | 14 | 13.1 |
| **No** | 93 | 86.9 |
| **Total** | 107 | 100 |
| History of thrombophilic condition |  |  |
| **yes** | 5 | 4.7 |
| **No** | 102 | 95.3 |
| **Total** | 107 | 100 |
| Active cancer |  |  |
| **No** | 98 | 91.6 |
| **Yes** | 9 | 8.4 |
| **Total** | 107 | 100 |
| Heart failure(s) |  |  |
| **No** | 98 | 91.6 |
| **Yes** | 9 | 8.4 |
| **Total** | 107 | 100 |
| Age in years |  |  |
| **<70** | 86 | 80.4 |
| **≥70** | 21 | 19.6 |
| **Total** | 107 | 100 |
| Ischemic stroke |  |  |
| **No** | 96 | 89.7 |
| **Yes** | 11 | 10.3 |
| **Total** | 107 | 100 |
| Acute infection |  |  |
| **No** | 76 | 71 |
| **Yes** | 31 | 29 |
| **Total** | 107 | 100 |

**Table 4.** Bivariate analysis of the leading diagnosis and Padua predictionscore Source: Authors’ Compilation, 2020

|  | PPS SCORE |  |  |  |
| --- | --- | --- | --- | --- |
|  | <4 score | ≥4 score | Total | P-value |
| Leading Diagnosis | Number (%) | Number n(%) | No. |  |
| DKA |  |  |  | 0.173 |
| **No** | 17(17.3) | 81(82.7) | 98 |  |
| **Yes** | 0(0) | 9(100) | 9 |  |
| **Total** | 17(15.9) | 90(84.1) | 107 |  |
| Malignancy |  |  |  | 0.173 |
| **No** | 17(17.3) | 81(82.7) | 98 |  |
| **Yes** | 0(00) | 9(100) | 9 |  |
| **Total** | 17(15.9) | 90(84.1) | 107 |  |
| Heart failure |  |  |  | 0.156 |
| **No** | 17(17.3) | 81(82.7) | 98 |  |
| **Yes** | 0(0) | 9(100) | 9 |  |
| **Total** | 17(15.9) | 90(84.1) | 107 |  |
| Complicated Malaria |  |  |  | 0.128 |
| **No** | 17(17.7) | 79(82.3) | 96 |  |
| **Yes** | 0(0) | 11(100) | 11 |  |
| **Total** | 17(15.9) | 90(84.1) | 107 |  |
| Severe Pneumonia |  |  |  | **0.004\*\*\*** |
| **No** | 17(22.4) | 59(77.6) | 76 |  |
| **Yes** | 0(0) | 31(100) | 31 |  |
| **Total** | 17(15.9) | 90(84.1) | 107 |  |
| EXTRA PULMONARY TB |  |  |  | 0.273 |
| **No** | 17(16.8) | 84(83.2) | 101 |  |
| **Yes** | 0(0) | 6(100) | 6 |  |
| **Total** | 17(15.9) | 90(84.1) | 107 |  |
| ISCHEMIC STROKE |  |  |  | 0.128 |
| **No** | 17(17.7) | 79(82.3) | 96 |  |
| **Yes** | 0(0) | 11(100) | 11 |  |
| **Total** | 17(15.9) | 90(84.1) | 107 |  |
| Meningitis |  |  |  | 0.234 |
| **No** | 17(17) | 83(83) | 100 |  |
| **Yes** | 0(0) | 7(100) | 7 |  |
| **Total** | 17(15.9) | 90(84.1) | 107 |  |
| Uncontrolled DM |  |  |  | **<0.001\*\*\*** |
| **No** | 5(5.4) | 88(94.6) | 93 |  |
| **Yes** | 12(85.7) | 2(14.3) | 14 |  |
| **Total** | 17(15.9) | 90(84.1) | 107 |  |

**Table 5.** Anticoagulant prophylaxis use in patients at VTE risk Source: Authors’ Compilation, 2020

|  | VTE prophylaxis status in VTE Risk patients |  |  |
| --- | --- | --- | --- |
| Leading Diagnosis | **Yes: numbers (%)** | **No:n(%)** | **P-value** |
| DKA |  |  | 0.694 |
| **No** | 6(7.4) | 75(92.6) |  |
| **Yes** | 1(11.1) | 8(88.9) |  |
| Malignancy |  |  | 0.694 |
| **No** | 6(7.4) | 75(92.6) |  |
| **Yes** | 1(11.1) | 8(88.9) |  |
| Heart failure |  |  | 0.694 |
| **No** | 6(7.4) | 75(92.6) |  |
| **Yes** | 1(11.1) | 8(88.9) |  |
| Complicated Malaria |  |  | 0.862 |
| **No** | 6(7.6) | 73(92.4) |  |
| **Yes** | 1(9.1) | 10(91) |  |
| Severe Pneumonia |  |  | 0.626 |
| **No** | 4(6.8) | 55(93.2) |  |
| **Yes** | 3(9.7) | 28(90.3) |  |
| EXTRA PULMONARY |  |  | 0.462 |
| **No** | 7(8.3) | 77(91.7) |  |
| **Yes** | 0(0.0) | 6(100) |  |
| ISCHEMIC STROKE |  |  | 0.862 |
| **No** | 6(7.6) | 73(92.4) |  |
| **Yes** | 1(9.1) | 10(91) |  |
| Meningitis |  |  | **0.032\*\*** |
| **No** | 5(6) | 78(94) |  |
| **Yes** | 2(28.6) | 5(71.4) |  |
| Uncontrolled DM |  |  | 0.678 |
| **yes** | 0(0.0) | 2(100) |  |
| **No** | 7(8) | 81(92) |  |

**Table 6.** Padua prediction score and VTE Prophylaxis Source: Authors’ Compilation, 2020

| Padua prediction score vs previous VTE Prophylaxis |  |  |  |
| --- | --- | --- | --- |
|  | <4 score | ≥4 score | P-value |
| **VTE prophylaxis** | n (%) | n (%) | 0.234 |
| **yes** | 0(0) | 7(100) |  |
| **No** | 17(17) | 83(83) |  |
| **Total** | 17(15.9) | 90(84.1) |  |

**Figure 1.**
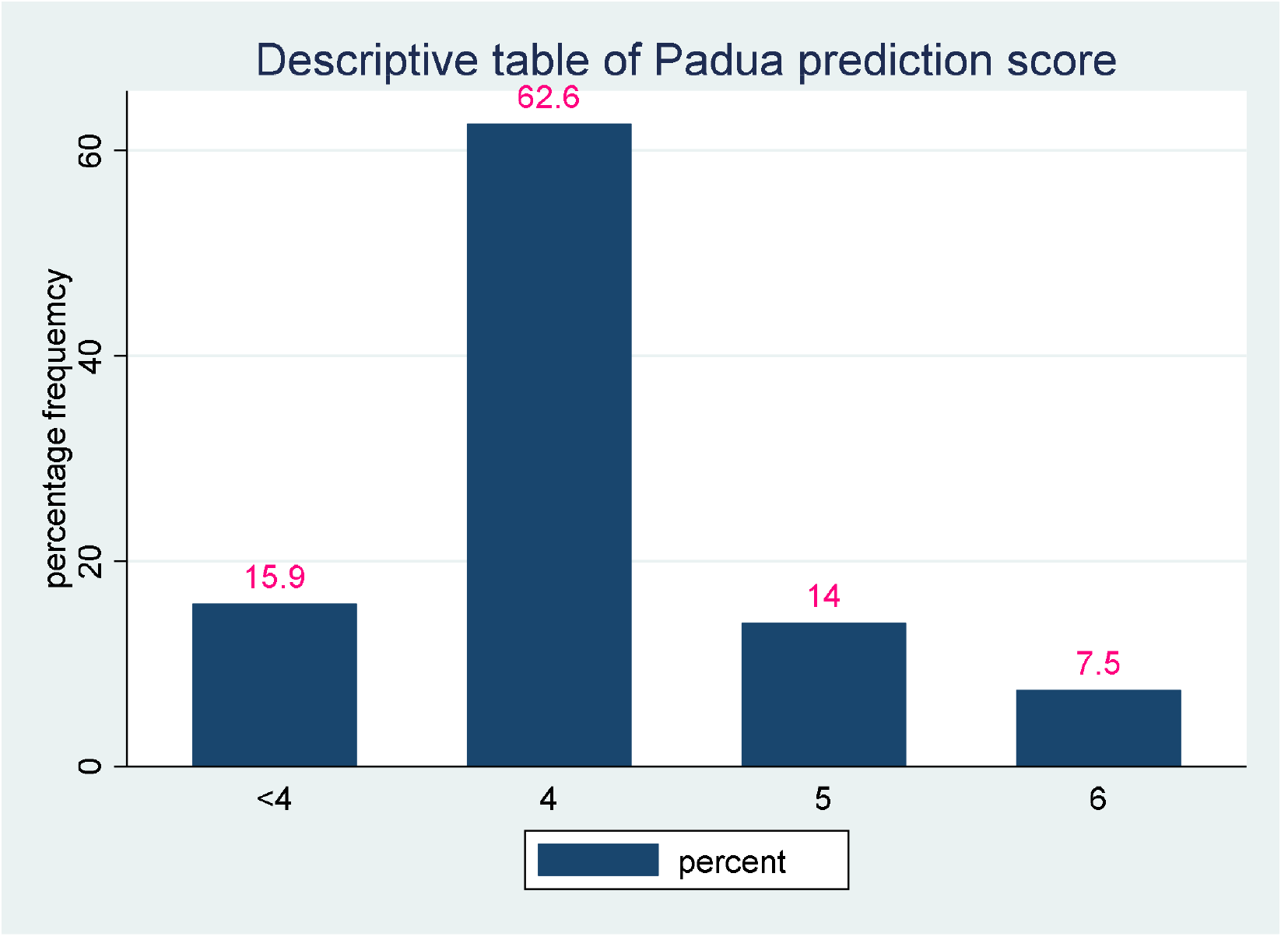
The distribution of Padua prediction score. Source: Authors’ Compilation, 2020

**Figure 2.**
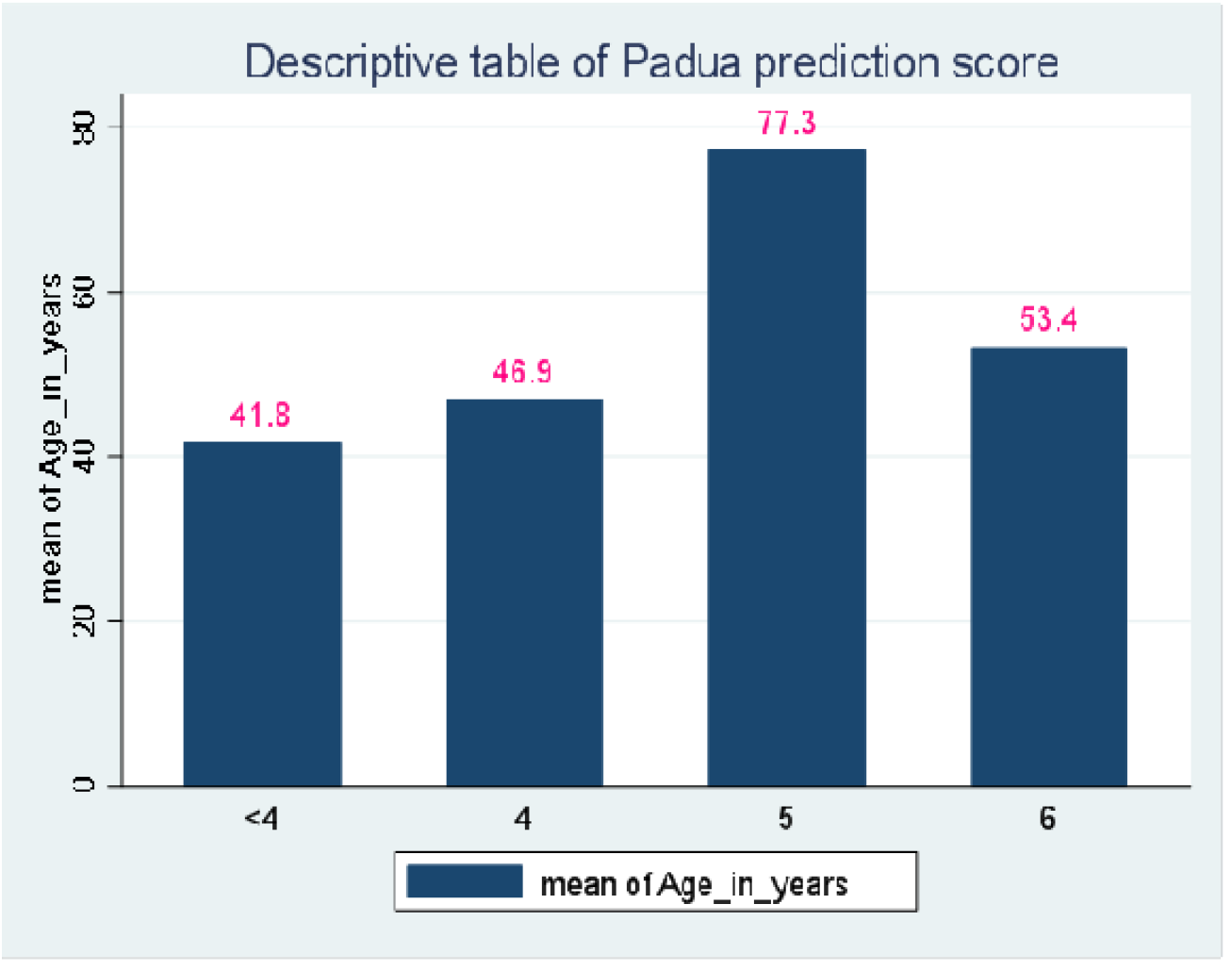
The disribution of Padua score by age in years Source: Authors’ Compilation, 2020

## DISCUSSION

VTE is a major problem in non-surgical hospitalized patients, needs more attention as it leads to substantial costs and it is associated with high morbidity and mortality [12]. Every year in the United States, VTE is diagnosed at the first time in 100 persons per 100,000, and its incidence is in rise from <5 cases per 100,000 persons in <15 years old to 500 cases per 100,000 persons at age of 80 years old [13]. The venous thrombophylaxis is extensively under-usage in hospitalized medical patients, a simple risk assessment model (RAM) was used to assess high risk VTE patients who received venous thromboprophylaxis in comparison with who did not, the findings, 2.2% who received VTE prophylaxis developed VTE and 11% in comparison group [9]. In the group of high risk VTE patients, the mean age of 77.7 years; reduced mobility; acute infection or rheumatologic disorder; and Heart failure and or respiratory failure showed significant association with VTE with p values <0.01 [9]; and the relative risk was 38.8 (95% CI, 10.4-146.5) [9]. The VTE free cumulative proportion was increasing with absence of VTE prophylaxis use and increased length of hospital stay [9]. The patients who scored ≥ 4 and did not receive prophylaxis had 30 times risk of developing in-hospital VTE and adequate thrombophylaxis reduced the risk at 90% [9]. Previous study conducted in two tertiary hospitals in Rwanda, the prevalence of proximal DVT was 5.5% in patients who were admitted in Internal Medicine and Obstetrics-Gynecology Departments, among them 75% were taking anticoagulant drugs and this study has documented low use of anticoagulant thromboprophylaxis [14].

Our findings in CHUK, the severely ill medical patients are not assessed properly and there was underuse of anticoagulant thromboprophylaxis to eligible patients. Among 107 recruited patients, 90 (84.1%) patients had VTE risk or they should be even many if we were able to assess BMI (Table 2) and among them only 10 (11.1%) patients were taking anticoagulant thromboprophylaxis (Figure 3). Among the patients who were physiologically unstable, 60.7% were classified in red color (Table 2), this is explained by referring system in public hospitals where the patients are getting the first treatment at health post or health center and if no improvement, they are referred at district hospital, referral hospital or provincial hospital then later to the Teaching Hospital, the patients may delay and their health conditions become worse. Among leading diagnosis, severe pneumonia was predominant followed by complicated malaria and ischemic stroke (Table 2). Four five of patients were aged below 70 years old, compared to the previous study where mean age was 77.7 years and showed significant association with VTE risk [9] contrary to our study, around 10 patients who scored 5 points were aged 78 years old (Figure 1, Figure 2). In this study, severe pneumonia and uncontrolled DM showed significant association with VTE risk (Table 4), here the study shared the same finding with previous study with one parameter of infection [9]. This study showed that aging was associated with increasing VTE risk where patients with mean age of 41.8 years old had no risk of VTE while the patients with mean age of 46.9 years old, 77.3 years old and 53.4 years old scored 4 points, 5 points and 6 points respectively (Figure 2). 62.6% of the patients who had mean aged of 46.9 years old scored 4 points (Figure 1, Figure 2), this was explained by young age is associated with high mobility and less Padua prediction score (9).

**Figure 3.**
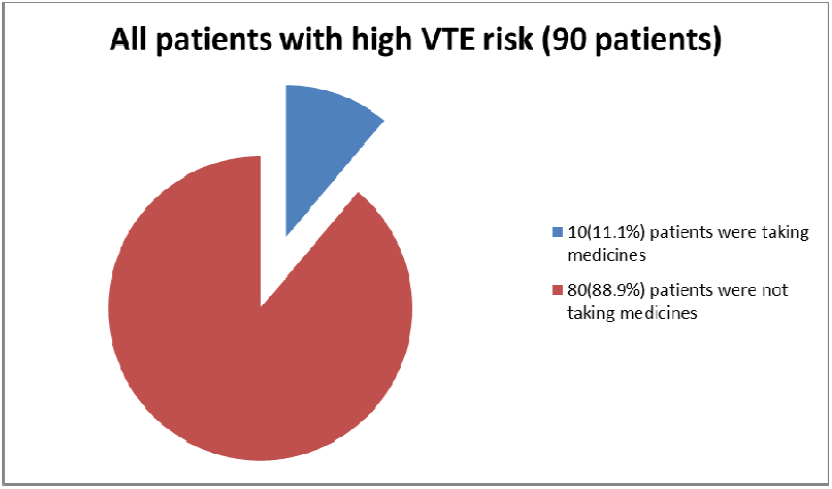
Pie Chart of all high VTE risk patients comparing the patients who were taking anticoagulant thromboprophylaxis and ones were not taking anticoagulant thromboprophylaxis prior to recruitment Source: Authors’ Compilation, 2020

## CONCLUSIONS AND RECOMMENDATIONS

Based on our study findings, there was high risk of developing VTE in admitted patients and underuse of anticoagulant thromboprophylaxis in potential candidates at CHUK. Padua prediction score should be implemented at the point of entry to assess high risk patients and start anticoagulant thrombophylaxis on time in non-surgical patients. At best of our knowledge, this is the first study assessing VTE risk and the use of anticoagulant thromboprophylaxis in CHUK, as well in Rwanda. Due to the lack of national standard protocol on use of anticoagulant thromboprophylaxis in severely ill non-surgical patients, the Padua prediction VTE risk assessment should be implemented to detect all patients at VTE risk among physiologically unstable patients as well the accessibility of anticoagulant thrombophylaxis in all hospitals in Rwanda to reduce mortality and morbidity.

Due to the time frame limitation and lack of external funds, we were not able to do follow-up of our patients during hospitalization and after discharge for assessing the treatment response for ones who received prophylaxis in terms of success rate.

## Data Availability

Dataset is safely maintained by investigators and may be shared on demand

## Acknowledgements

We acknowledge all clinical staff of CHUK, especially Accident and Emergency staff and Internal medicine ward, and all patients who accepted to participated in this study

## Authors’ contribution

**EA** and **EN** were responsible for literature search and drafting of the article, **EA, EN** and **RN** were responsible for conception of article; and **EN, OS** and **FM** were responsible for revision of article.

## Ethics approval

This study was approved by Institutional Review Board of College of Medicine and Health Sciences (No 032/CMHS/IRB2020) and Ethics committee of CHUK (Ref: EC/CHUK/017/2020). Prior to data collection, an informed consent was obtained from patients or next of kin (who is legally accepted) in case the patients are not able to give his/her consent.

## Data availability statement

The dataset is well maintained by the study investigators and can be shared on demand.

